# Comparison of the coronavirus pandemic dynamics in Europe, USA and South Korea

**DOI:** 10.1101/2020.03.18.20038133

**Authors:** Igor Nesteruk

## Abstract

The pandemic cased by coronavirus COVID-19 is of great concern. A detailed scientific analysis of this phenomenon is still to come, but now it is urgently needed to evaluate the disease dynamics in order to organize the appropriate quarantine activities, to estimate the required number of places in hospitals, the level of individual protection, the rate of isolation of infected persons, etc. South Korea has achieved the stabilization of the number of cases at rather low level. The epidemic dynamics there can be compared with its development in other countries to make some preliminary, but very important conclusions. Here we provide a simple method of data comparison that can be useful for both governmental organizations and anyone.

## Introduction

Some previous efforts to compare the epidemic dynamics in Italy and mainland China has been done in [1, 2]. Here we will try to compare the epidemics in Italy, Spain, Germany, France, Switzerland, and USA with the situation in South Korea, where the number of cases is stabilizing at rather low level and the mortality rate is not high. We improved the very simple method proposed in

[1] which now can by applied for every country. To illustrate its application, the epidemic dynamics in Austria was estimated.

### Data

We will use official data about the accumulated number of confirmed cases in Italy, Spain, France, Germany, Switzerland, USA, South Korea *V*_*j*_ from WHO daily situation reports [3], see

Tables 1 and 2. Since the reports show the numbers accumulated by 10AM CET, we assumed that every number *V*_*j*_ corresponds to the previous day. The values *V*_*j*_ and corresponding moments of time *t*_*j*_ and *t*_*ej*_ are shown in Tables 1 and 2.

**Table 1.** Official cumulative numbers of confirmed cases in the Republic of Korea. The information from [3]. The corresponding time moments *t*_*j*_ and the accumulated confirmed numbers of cases *V*_*j*_ in South Korea.

| Day in<br>February,<br>2020 | Time<br>moment<br>$t_j$ | Accumulated<br>number of cases<br>in the Republic<br>of Korea $V_j$, [3] | Day in<br>March,<br>2020 | Time<br>moment<br>$t_j$ | Accumulated<br>number of cases<br>in the Republic of<br>Korea $V_j$, [3] |
| --- | --- | --- | --- | --- | --- |
| 17 | -1 | 31 | 1 | 12 | 4212 |
| 18 | 0 | 51 | 2 | 13 | 4812 |
| 19 | 1 | 104 | 3 | 14 | 5328 |
| 20 | 2 | 204 | 4 | 15 | 5766 |
| 21 | 3 | 346 | 5 | 16 | 6284 |
| 22 | 4 | 602 | 6 | 17 | 6767 |
| 23 | 5 | 763 | 7 | 18 | 7134 |
| 24 | 6 | 977 | 8 | 19 | 7382 |
| 25 | 7 | 1261 | 9 | 20 | 7513 |
| 26 | 8 | 1766 | 10 | 21 | 7755 |
| 27 | 9 | 2337 | 11 | 22 | 7869 |
| 28 | 10 | 3150 | 12 | 23 | 7979 |
| 29 | 11 | 3736 | 13 | 24 | 8086 |
| - | - | - | 14 | 25 | 8165 |

**Table 2.** The number of cases in Italy, Spain, France, Germany, Switzerland and USA used for comparisons and the results of calculations. The information about the confirmed accumulated number of cases from [3]. The mortality rate was calculated with the use of numbers for March 14, 2020 (the situation report dated March 16, 2020, [3]). The corresponding time moments *t*_*ej*_ are shown in the last column. The values corresponding the beginning of the epidemic outbreak are shown in red. The values *d*_*t*_ were calculated with the use of formula (2).

| Day in February<br>and March, 2020 | Italy | Germany | France | Spain | Switzerland | USA | $t_{ej}$ |
| --- | --- | --- | --- | --- | --- | --- | --- |
| 22 | 76 | 16 | 12 | 2 | 0 | 35 | 0 |
| 23 | 124 | 16 | 12 | 2 | 0 | 35 | 1 |
| 24 | 229 | 16 | 12 | 2 | 0 | 53 | 2 |
| 25 | 322 | 18 | 12 | 2 | 1 | 53 | 3 |
| 26 | 400 | 21 | 18 | 12 | 1 | 59 | 4 |
| 27 | 650 | 26 | 38 | 25 | 6 | 59 | 5 |
| 28 | 888 | 57 | 57 | 32 | 10 | 62 | 6 |
| 29 | 1128 | 100 | 57 | 45 | 18 | 62 | 7 |
| 1 | 1689 | 129 | 100 | 45 | 24 | 62 | 8 |
| 2 | 2036 | 157 | 191 | 114 | 30 | 64 | 9 |
| 3 | 2502 | 196 | 212 | 151 | 37 | 108 | 10 |
| 4 | 3089 | 262 | 282 | 198 | 56 | 129 | 11 |
| 5 | 3858 | 534 | 420 | 257 | 86 | 148 | 12 |
| 6 | 4636 | 639 | 613 | 374 | 209 | 213 | 13 |
| 7 | 5883 | 795 | 706 | 430 | 264 | 213 | 14 |
| 8 | 7375 | 1112 | 1116 | 589 | 332 | 213 | 15 |
| 9 | 9172 | 1139 | 1402 | 1024 | 332 | 472 | 16 |
| 10 | 10149 | 1296 | 1774 | 1639 | 491 | 696 | 17 |
| 11 | 12462 | 1567 | 2269 | 2140 | 645 | 987 | 18 |
| 12 | 15113 | 2369 | 2860 | 2965 | 858 | 1264 | 19 |
| 13 | 17660 | 3062 | 3640 | 4231 | 1125 | 1678 | 20 |
| 14 | 21151 | 3795 | 4469 | 5753 | 1359 | 1678 | 21 |
| Mortality rate (%),<br>March 14, 2020 | 6.81 | 0.21 | 2.04 | 2.36 | 0.81 | 2.44 | - |
| $d_t$ | 0.549 | 0.154 | 0.154 | -0.179 | 0.129 | 0.312 | - |

### Time synchronization procedure

We have to calculate the number of days from start of the epidemic outbreak (which is different for every country, see red values in the last column of Table 2) to compare with the data set listed in Table 1. To increase the accuracy, we let this number to be non-integer. To calculate the corresponding time difference *d*_*t*_, we use the parabolic interpolation for the initial number of cases *V* in South Korea. The first three points in Table 1 yield the corresponding equation:

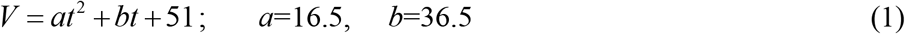

Then by putting into (1) the values *V=V*_*b*_ for the number of cases at the starting day of the epidemic outbreak (shown in red in Table 2), the corresponding values *d*_*t*_ were calculated for every country with the use of formula

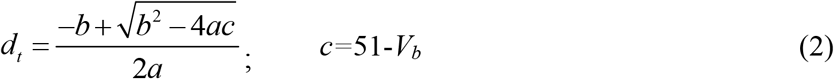

which yields a solution to the quadratic equation (1). The calculated values of *d*_*t*_ are shown in the last row of Table 2. Then the time moments for Italy have to be shifted by 0.549 days, for Germany by 6+0.154 days and so on.

## Results and discussion

The synchronized data sets are shown in the Fig. 1 by “stars” for countries in Europe and by “squares” for USA. The data for South Korea are shown by red line with “circles”. Blue “stars” represent the sum of cases for 5 countries in Europe. It can be seen that Italy, Spain and France have no chance for rapid stabilization of the number of cases. But it is still possible in Germany, Switzerland and USA. Very high mortality rate in Italy and Spain makes the situation really dramatic. Blue “stars” in the Fig. 1 still follow the straight line. It means that the number of cases in Europe still increases exponentially and is far from stabilization.

**Fig 1.**
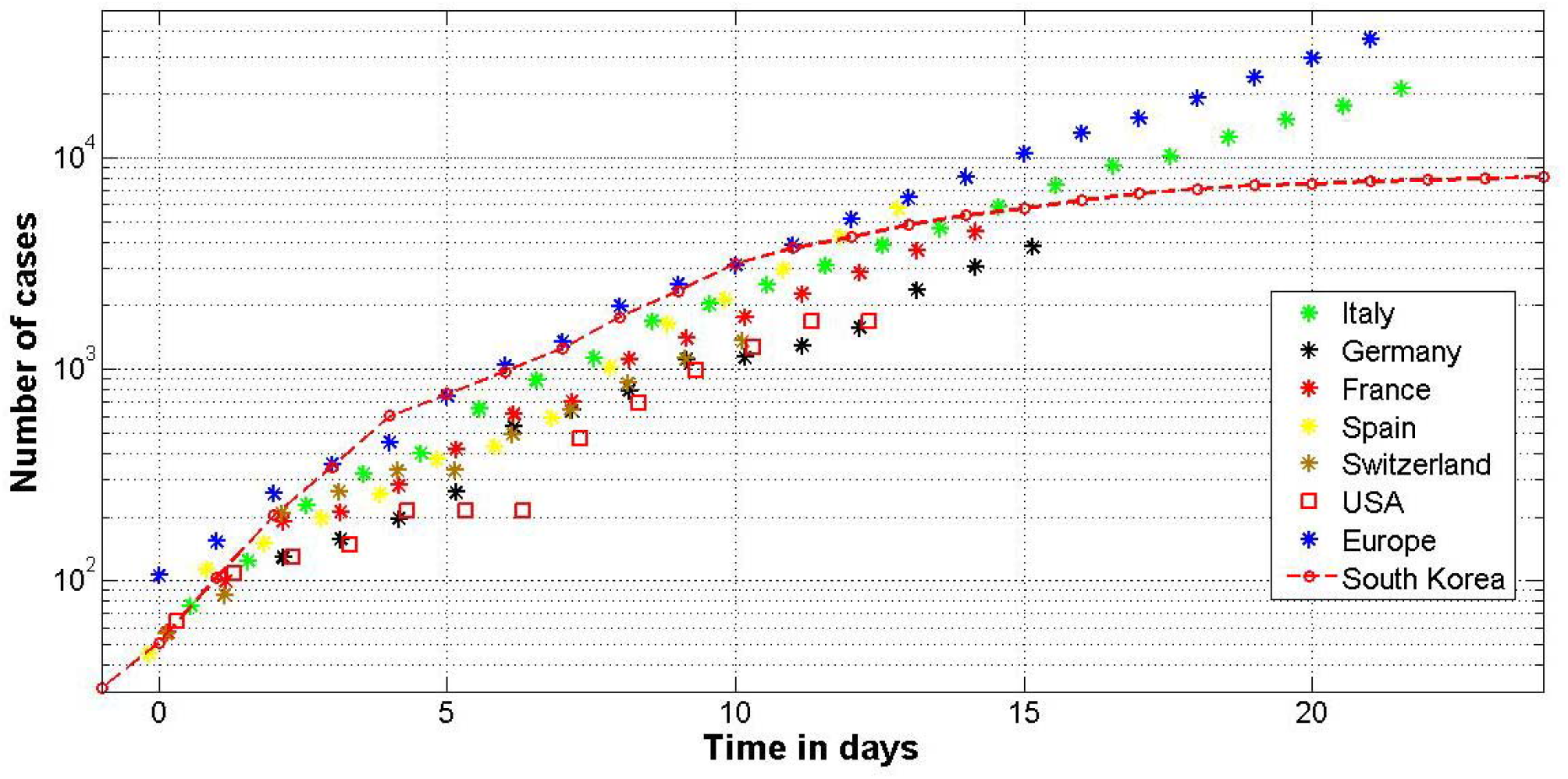
The synchronized data sets for the accumulated confirmed number of cases versus time in days from the beginning of the epidemic outbreaks.

It seems that main reason for the rapid increase in the number of cases in Italy and Spain is the slow isolation of infected and contact persons. For example, in South Korea an infected person spreads the infection approximately 4.3 hours (in average), [4]; the mortality rate is 0.92%. By comparison, in mainland China this time is estimated as 2.5 days [5].

### How to estimate the epidemic dynamics for other countries?

For example, let us illustrate the application of this simple procedure for Austria. The number of cases in this country was *V*_*b*_ =66 on March 6, 2020. Let us suppose that epidemic in this country started on this day. Corresponding value of *t*_*ej*_ = 13 (see Table 2) and *d*_*t*_ =0.3542 (according to eq. (2)). If we want to know how looks the situation on March 15, 2020 (ninth day of the epidemic), we open the corresponding WHO report (number 56) and find their the number 959 for Austria. This figure has to be compared with the number of cases from Table 1, corresponding to the time moment 9+0.3542. To avoid any additional calculations, let us take the value 2337 for comparison. Thus the situation in Austria does not look bad. This country is able to stabilize the number of cases soon.

## Conclusions

The situation with the coronavirus pandemic in Europe is very threatening. Italy and Spain are in urgent need of assistance in speeding up the isolation of infected and contact persons.

## Data Availability

All data are in the text

## Acknowledgements

I would like to express my sincere thanks to Ihor Kudybyn for his help in collecting and processing data.

